# Can neuromuscular electrostimulation fulfil an unmet need for mechanical thromboprophylaxis in acute stroke patients? A real-world retrospective analysis

**DOI:** 10.1101/2025.06.10.25329340

**Authors:** Indira Natarajan, Alison Griffin, Lakshmanan Sekaran, Sakthivel Sethuraman, Jane Sword

## Abstract

**Aims and Method:** This retrospective analysis examines real-world registry data to assess the incidence of symptomatic VTE in a non-randomised cohort where IPC was either contraindicated or not tolerated, leading to the use of Neuromuscular electrostimulation (NMES) to meet prophylactic needs. We consider the alternative VTE risk of no prophylactic intervention other than standard measures and utilise the data from the CLOTS 3 trial to evaluate the incidence of symptomatic VTE in a cohort that was randomly assigned to receive no IPC intervention, The outcomes from both cohorts are then compared to determine which group exhibits better clinical results.

**Results:** A comparison of symptomatic VTE incidence revealed a significantly lower rate in patients prescribed NMES as their first-line intervention of 1.5% compared to no IPC reported in CLOTS 3 (8.7%). The reduction in VTE risk between no IPC treatment and NMES as prescribed as a front-line sole intervention in the unmet need is statistically significant. (p<0.00001, Fisher’s exact test).

**Health Economics:** In terms of an economic analysis the average treatment with the NMES device in this pathway is 9 days and the cost of NMES is £243. In the unmet need scenario described above this results in an approximate 5% reduction in symptomatic VTE risk compared to no IPC intervention. The economic saving of this reduction in VTE risk compared to no IPC is modelled to be £303 per patient.

**Conclusion:** This retrospective analysis indicates that NMES offers an effective alternative for VTE mechanical prophylaxis in immobile acute stroke patients, a high-risk cohort, delivering significantly improved VTE outcomes compared to no IPC treatment. Further clinical insight from this registry suggests that the VTE outcomes achieved with the NMES device are comparable to those reported for IPC, further supporting its inclusion in the VTE prevention pathway.

## Introduction

### Risk

Stroke patients are at high risk of venous thromboembolism (VTE) ^i^, which includes both deep vein thrombosis (DVT) and pulmonary embolism (PE). The risk of VTE peaks within the first two weeks following a stroke^ii^ and remains elevated for several months, with evidence suggesting that the risk may even precede the stroke event^iii^. Among stroke patients, DVT specifically is an independent predictor of poor prognosis and mortality within three months^iv^.

After the stroke itself and secondary infections, VTE is one of the leading causes of death in stroke patients^v vi^, contributing to 13%-25% of post-stroke fatalities^vii^. While VTE is considered a preventable complication ^viii^, the implementation of thromboprophylactic measures has become a key quality indicator in stroke care centres^ix^.

Over the past four decades, advancements in post-stroke care and VTE management have significantly reduced the reported incidence of VTE ^x^. However, interpreting these data remains challenging due to variations in the timing and methods of screening.

Earlier studies conducted before routine prophylaxis reported VTE rates (which include both DVT and PE) ranging from 40%-60% ^xi xii^ using methods such as 125I-fibrinogen scanning. More recent studies report VTE incidences of 20%-50% with similar methods, 7%-20% by Doppler ultrasound, and approximately 15%-25% using magnetic resonance direct thrombus imaging. Whereas symptomatic VTE rates (which include both DVT and PE) typically range from 2%-10% in stroke patients ^viii xiii^.

### Prophylaxis and Unmet Need

Thromboprophylactic measures that have been confirmed effective through randomised controlled trials (RCTs) and systematic reviews include both pharmacologic and mechanical interventions. It is well-established that anticoagulant therapy, such as low molecular weight heparin, effectively reduces VTE incidence. ^xiv^ However, its use is associated with an increased risk of bleeding, and guidelines recommend against its use, particularly in patients with haemorrhagic stroke. ^xv^

NICE guidance NG 89 recommends Intermittent Pneumatic Compression (IPC) for acute stroke due to its proven effectiveness in reducing VTE risk without increasing the risk of haemorrhage. ^xvi^ IPC reduces venous stasis by applying intermittent compression to the legs ^xvi^, which has been shown to decrease the overall incidence of VTE in acute stroke by approximately 2.5%–5% in high-risk populations. When IPC is combined with pharmacological prophylaxis, the risk of VTE is reduced even further, offering enhanced protection against both DVT and PE^xvii, xviii^. However, current UK guidelines do not recommend chemical prophylaxis in acute stroke patients due to risk of further haemorrhage complications.^xviii^ It should also be noted that IPC is contraindicated in patients with conditions such as dermatitis, leg ulcers, severe oedema, peripheral vascular disease, and congestive heart failure. ^xix^ Additionally, optimum fitting and adherence to IPC remains a challenge, ranging from 40% to 89%, ^xx^ with common barriers including discomfort, difficulty with ambulation, noise, and machine malfunction. ^xxi^

These contraindications and adherence challenges represent a significant patient population whose prophylactic needs remain unmet.

### Neuromuscular Electrostimulation (NMES) of the Common Peroneal Nerve

The National Institute for Healthcare and Excellence (NICE) guidance MTG19^xxii^ recommend the use of the geko® device, a neuromuscular electro stimulator (NMES) for hospitalised patients who have a high-risk of VTE and for whom other mechanical and pharmacological methods of prophylaxis are impractical or contraindicated.^xxiii^

The geko® device is a simple, button-cell-powered, wearable device that delivers intermittent (1Hz) transdermal NMES to the common peroneal nerve, resulting in cyclic muscle contractions in the leg. These contractions activate the venous valve pumps in the leg and foot, ^xxiv^ ^xxv^ helping to reduce venous stasis, increase shear forces on the endothelial lining, which reduces coagulability, and mobilise the valves, alleviating stasis in the valve sinuses. ^xxvi^ The device is self-adhesive, discreet, and wire-free, making it easy to apply either by the clinician or the patient (see Figure 1).

**Figure 1.**
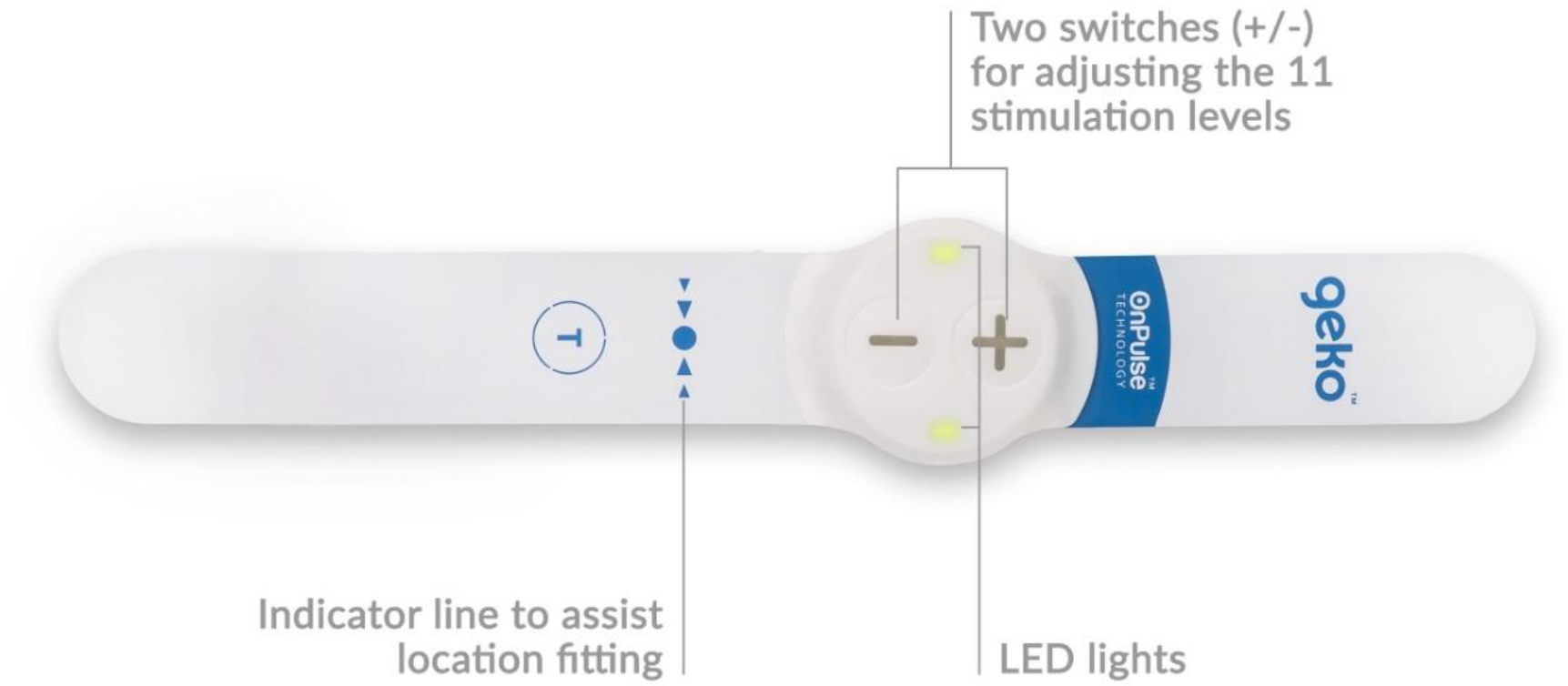
The geko® device, Firstkind Ltd, Daresbury.

**Figure 2.**
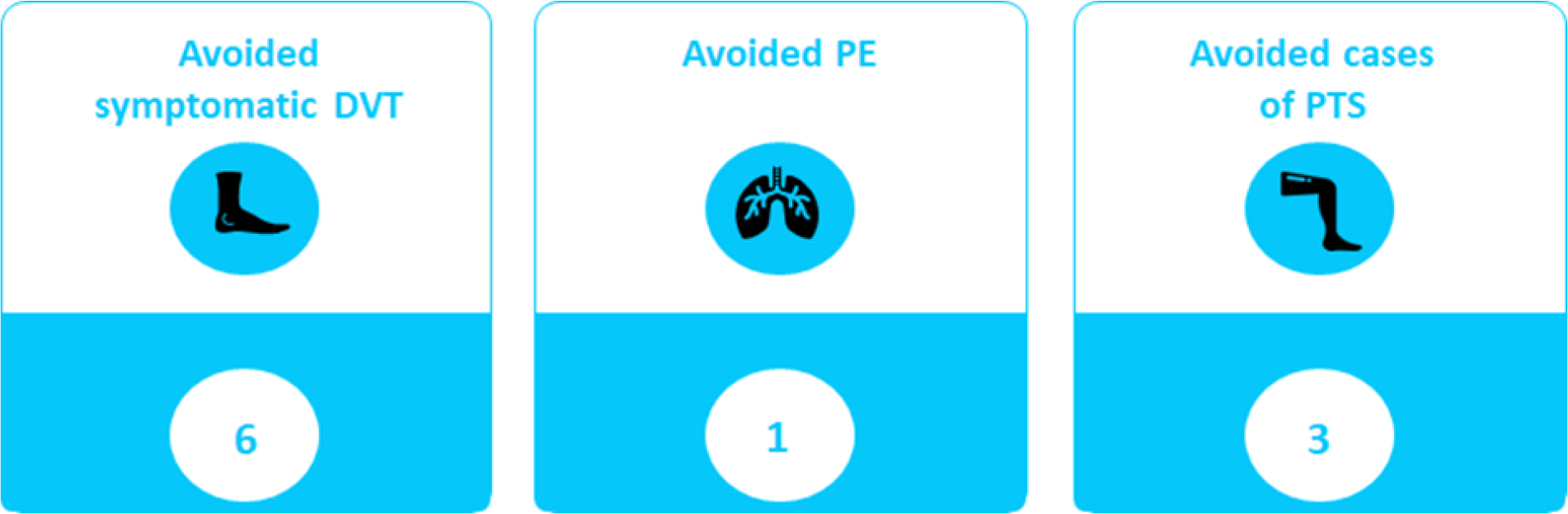
illustrates the rate of events avoided per 100 patients.

NMES of the common peroneal nerve has been shown to increase both venous volume and velocity in the lower limb. ^xxvii^ ^xxviii^ ^xxix xxx^ Doppler ultrasound measurements of deep veins demonstrate substantial increases in peak velocity and ejected volume in the peroneal, posterior tibial, and gastrocnemius veins. ^xxxi^ Compared to IPC, NMES has been shown to be superior in enhancing blood flow in the lower limbs, while also being safe and well-tolerated by patients. ^xxxii^ ^xxxiii^

In addition to improving venous circulation, NMES reduces venous transit time, ambulatory venous pressure, and leg volumes, and increases muscle blood oxygenation.^xxxiv xxxv^ However, no prospective RCT has yet been conducted to compare the effectiveness of NMES with other prophylactic measures in the stroke population. Given the sporadic nature of DVT incidence, such a study would require a large sample size, potentially involving thousands of participants, to achieve statistical significance.

### Aims

This retrospective analysis examines real-world registry data to assess the incidence of symptomatic VTE in a non-randomised cohort where IPC was either contraindicated or not tolerated, leading to the use of NMES to meet prophylactic needs. We consider the alternative VTE risk of no prophylactic intervention other than standard measures and utilise the data from the CLOTS 3 trial to evaluate the incidence of symptomatic VTE in a cohort that was randomly assigned to receive no IPC intervention, The outcomes from both cohorts are then compared to determine which group exhibits better clinical results.

An economic evaluation is then conducted using the cost-consequence economic model developed for the NICE Medical Technologies Guidance 19 (MTG19). This model assesses the financial benefits of using the NMES device compared to the cost consequence of no intervention in the described scenario of unmet need.

Unlike cost-effectiveness analyses, which generate a single ratio combining both quantity and quality of life gained from a treatment (cost per QUALY), a cost-consequence model separately presents costs (e.g., device expenses, healthcare savings) and consequences (e.g., reduced DVT incidence, improved patient outcomes).

This approach aims to demonstrate that the use of the NMES device will provide both clinical and economic benefits in a scenario of unmet prophylactic need.

### Analysis Datasets

#### 1 Real-World Registry Data

Real-world registry data, collected by the Acute Stroke Services at three UK hospitals and considered suitable for evaluation, were obtained from the sources summarised in Table 1.

**Table 1:**
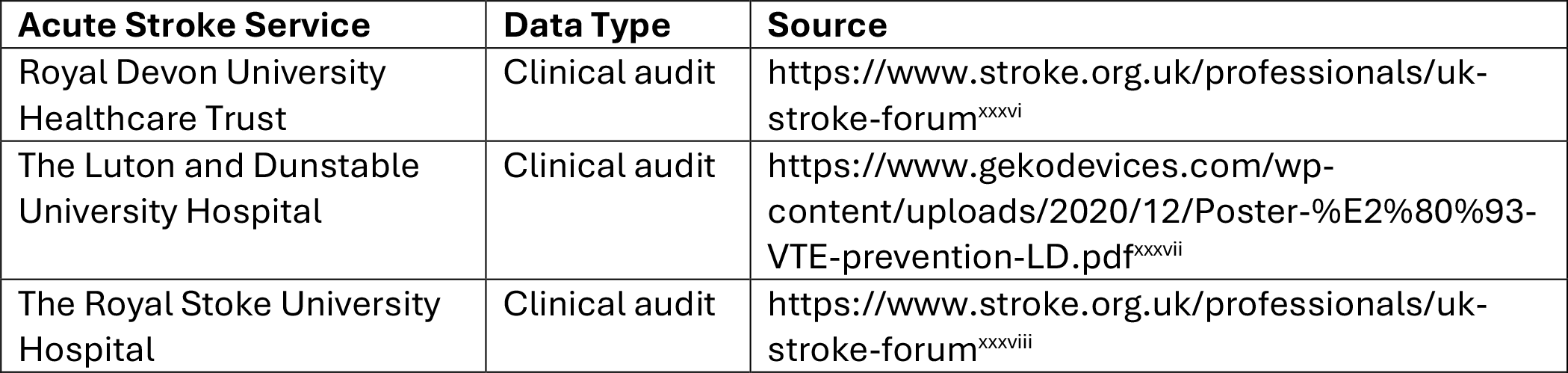
Real-World Registry Data Sources.

**Table 2:** Summary of Real-World Registry Data from Patients Reviewed at three UK Hospitals

|  | Patients reviewed | IPC only | IPC then NMES (IPC intolerant) | NMES only | IPC + anti-coagulation | Other* |
| --- | --- | --- | --- | --- | --- | --- |
| The Royal Devon University Healthcare NHS Trust | 180 | 81 | 11 | 1 | 0 | 87 |
| The Royal Stoke University Hospital | 2000 | 805 | 156 | 455 | 0 | 584 |
| The Luton and Dunstable University Hospital | 215 | 128 | 0 | 12 | 75 | 0 |
| **TOTAL** | **2395** | **1014** | **167** | **468** | **75** | **671** |

\*Prophylaxis not required (e.g. patient mobile, palliative care), anticoagulation given for other indications, non-concordance with treatment plan,

A total of 2,395 acute stroke patients were reviewed across three services. Of these, 1,724 patients (72%) were prescribed a form of mechanical prophylaxis, specifically IPC or NMES using the geko® device.

The 1724 patients prescribed mechanical prophylaxis, IPC was the primary treatment for 1,014 patients (59%) to meet their prophylactic needs. However, 635 patients (37%) who would have otherwise received IPC were either unable to be prescribed it or could not tolerate it, revealing a significant real-world unmet need. To address this, the NMES device was used as a first-line treatment in 468 of these 635 patients, covering 74% of the unmet need. For the remaining 26%, it served as a secondary intervention when IPC was not tolerated.

The remaining 671 patients (28%) were considered sufficiently mobile and did not require additional prophylaxis, were fully anticoagulated for other medical reasons or were receiving palliative care. This group also includes patients who could not tolerate any form of intervention.

#### 2. CLOTS 3 Trial Data

The CLOTS 3 trial was a multicentre, parallel-group study conducted across 105 hospitals in the UK. Patients were randomly assigned to receive either IPC or no IPC.

For this analysis, data from the ‘no IPC’ group from this trial has been used for a comparative evaluation or the potential risk associated with the unmet need described.

A summary of the data analysed is provided in Table 3.

**Table 3:** Summary of Data obtained from the CLOTS 3 Trial conducted across 105 UK Hospitals.

|  | Patients reviewed | IPC only | No IPC |
| --- | --- | --- | --- |
| CLOTS 3 Trial | 2876 | 1438 | 1438 |

## Results

### 1. Incidence of Symptomatic VTE

The symptomatic VTE risk outcomes for patients are summarised in Table 4. The real-world registry data set shows that for patients prescribed NMES as their sole first-line intervention, the incidence of symptomatic VTE was 1.5% (7/468). Whereas the symptomatic VTE risk outcomes for patients CLOTS 3 randomised to the group without IPC, representing an unmet prophylactic need of this registry data review, with 1,438 patients, experienced 125 cases of symptomatic VTE, yielding a risk outcome of 8.7%.

**Table 4:** Real-World Registry and CLOTS 3 Datasets: Symptomatic VTE Risk Outcome

| Source | Intervention | Number of Patients | Number of Symptomatic VTE Cases | Symptomatic VTE Risk Outcome (%) |
| --- | --- | --- | --- | --- |
| Real-World Registry Dataset | NMES (1st line intervention for patients with unmet prophylactic need) | 468 | 7 | 1.5 |
| CLOTS 3 Trial Data | No IPC (baseline risk associated with the unmet prophylactic need) | 1,438 | 125 | 8.7 |

### 2. Real-World Registry Data vs CLOTS 3 Trial Data

A comparison of symptomatic VTE incidence revealed a significantly lower rate in patients prescribed NMES as their first-line intervention of 1.5% compared to no IPC reported in CLOTS 3 (8.7%)

The reduction in VTE risk between no IPC treatment and NMES as prescribed as a front-line sole intervention in the unmet need is statistically significant. (p<0.00001, Fisher’s exact test).

### 3. Economic Impact of NMES

The real-world economic impact of using the NMES device in scenarios involving unmet prophylactic needs has been modelled using three key inputs: the average duration of device utilisation (9 days), the baseline risk of symptomatic VTE in immobile acute stroke patients unable to receive IPC (8.7%, comprising 6.3% DVT and 2.4% PE, as per the CLOTS 3 trial dataset), and the average VTE incidence in patients prescribed NMES as their first-line intervention (1.5%, comprising 0.2% DVT and 1.3% PE, based on the Real-World Registry dataset).

The cost of a symptomatic VTE as referenced in 2024 by NHS England as is £9722, the cost of a DVT event £2936, and the cost of a PE event is £6786, and the 15 year cost of post-thrombotic syndrome is reported to be £12088.

In terms of an economic analysis the average treatment with the NMES device in this pathway is 9 days and the cost of NMES is £243. In the unmet need scenario described above this results in an approximate 5% reduction in symptomatic VTE risk compared to no IPC intervention.

The economic saving of this reduction in VTE risk compared to no IPC is modelled to be £303 per patient.

## Discussion

This review of available real-world data indicates that the NMES device is a viable option for the significant proportion of patients (37%) whose needs are unmet by IPC and who would otherwise have no NICE approved mechanical prophylaxis available to them. Stroke patients prescribed NMES demonstrate a significantly lower incidence of symptomatic VTE compared to the reported risk of no IPC intervention, highlighting its real-world effectiveness. The NMES device is considered safe and has been positively received by nursing teams, providing an additional option for patients in this cohort and its integration into the pathways is described as straightforward. Furthermore, a health economic analysis based on 9 days of device use suggests that its implementation is cost-saving in this circumstance compared to the financial consequences of no IPC treatment. The cash value released suggests that the NMES device can be funded from both the saving in VTE events and the switching of consumable funds that would otherwise be allocated to patients for IPC.

## Conclusion

This retrospective analysis indicates that NMES offers an effective alternative for VTE mechanical prophylaxis in immobile acute stroke patients, a high-risk cohort, delivering significantly improved VTE outcomes compared to no IPC treatment. Further clinical insight from this registry suggests that the VTE outcomes achieved with the NMES device are comparable to those reported for IPC, further supporting its inclusion in the VTE prevention pathway. In summary, the NMES device is both clinically and economically beneficial for this cohort, strongly underpinning the NICE guidance recommendation.

## Data Availability

All data produced are available online at https://www.stroke.org.uk/professionals/uk-stroke-forum
https://www.gekodevices.com/wp-content/uploads/2020/12/Poster-%E2%80%93-VTE-prevention-LD.pdf 

## Notes

### Competing Interest Statement

The authors have declared no competing interest.

### Funding Statement

This study did not receive any funding its a retrospective analysis of data

### Author Declarations

The study used only openly available human data that were originally located at: Acute Stroke Service Data TypeSource Royal Devon University Healthcare TrustClinical audithttps://www.stroke.org.uk/professionals/uk-stroke-forum The Luton and Dunstable University HospitalClinical audit https://www.gekodevices.com/wp-content/uploads/2020/12/Poster-%E2%80%93-VTE-prevention-LD.pdf The Royal Stoke University HospitalClinical audithttps://www.stroke.org.uk/professionals/uk-stroke-forum

